# Revisiting prostate segmentation in magnetic resonance imaging (MRI): On model transferability, degradation and PI-RADS adherence

**DOI:** 10.1101/2023.08.21.23294376

**Authors:** Alvaro Fernandez-Quilez, Tobias Nordström, Trygve Eftestøl, Andreas Bremset Alvestad, Fredrik Jäderling, Svein Reidar Kjosavik, Martin Eklund

## Abstract

**Purpose:** To investigate the effect of scanner and prostate MRI acquisition characteristics when compared to PI-RADSv2.1 technical standards in the performance of a deep learning prostate segmentation model trained with data from one center (INST1), longitudinally evaluated at the same institution and when transferred to other institutions.

**Materials and Methods:** In this retrospective study, a nn-UNet for prostate MRI segmentation was trained with data from 204 patients from one institution (INST1) (0.50mm^2^ in-plane, 3.6mm thickness and 16cm field of view [FOV]). Post-deployment performance at INST1 was tested with 30 patients acquired with a different protocol and in a different period of time (0.60mm^2^ in-plane, 4.0mm thickness and 19cm FOV). Transferability was tested on 248 patient sequences from five institutions (INST2, INST3, INST4, INST5 and INST6) acquired with different scanners and with heterogeneous degrees of PI-RADS v2.1 technical adherence. Performance was assessed using Dice Score Coefficient, Hausdorff Distance, Absolute Boundary Distance and Relative Volume Difference.

**Results:** The model presented a significant degradation for the whole gland (WG) in the presence of a change of acquisition protocol at INST1 (DSC:99.46±0.12% and 91.24±3.32%,*P*<.001; RVD:-0.006±0.127% and 8.10±8.16%, *P*<.001). The model had a significantly higher performance in centers adhering to PI-RADS v2.1 when compared to those that did not (DSC: 86.24±9.67% and 74.83±15.45%, *P* <.001; RVD: -6.50±18.46% and 1.64±29.12%, *P*=.003).

**Conclusions:** Adherence to PI-RADSv2.1 technical standards benefits inter-institutional transferability of a deep learning prostate segmentation model. Post-deployment evaluations are critical to ensure model performance is maintained over time in the presence of protocol acquisition modifications.

## Introduction

Over the last decade, multi-parametric MRI (mp-MRI) has evolved as a key component for Prostate Cancer (PCa) detection, staging and treatment planning [1]. Its recommended upfront role and increasing relevance for PCa is expected to substantially increase the radiologist workload [2]. Delineation of the whole gland (WG) of the prostate plays an important role in PCa diagnosis and management, with important measures such as the volume being highly dependent on the quality and accuracy of it [3]. Further, reported markers of PCa risk such as PSA density (PSAd) are dependent on its accurate characterization, with mistakes potentially leading to an increase in patients recommended to undergo unnecessary biopsies and an increase in false positives in PCa detection [4, 5].

Existing shortcomings of manual MRI segmentation of the prostate WG in the form of inter and intra-reader variability [6] coupled with the growing shortage of available specialists, has motivated the development of a considerable amount of automatic prostate WG segmentation tools [1, 7]. Among the existing architectures, nn-UNet is commonly a top performer in prostate segmentation challenges and is considered as the *de facto* choice for the task [8, 9]. Whilst a broad range of results have been presented for prostate WG segmentation trained on *open-source* and *single-institution dataset*s [9, 10], little is known about the impact of the MRI scanner characteristics and their degree of adherence to PI-RADS v2.1 technical standards on the inter-institutional *transferability* and *longitudinal performance* of the model after a successful deployment [11]. Ultimately, those concerns have been shown to lead to issues on model adoption, usability and to obsoletion outside of research environments [12, 13, 14].

In this work, we aimed to (*a*) evaluate the *transferability* of a nn-UNet [15] prostate segmentation model without *calibration* (re-training) (*b*) evaluate the *temporal degradation* of the model and (*c*) investigate the effect of *technical acquisition and scanner characteristics* in the model performance in both scenarios. For that purpose, (*a*) we train the model on a *widely used and adopted* data cohort from one institution (INST1) [9] and evaluate it in five different institutions (INST2, INST3, INST4, INST5 and INST6) (*b*) on data from the same institution (INST1) acquired in a different period of time and with a different technical acquisition than the one used to train the model and (*c*) present a systematic characterization of the scanners and MRI technical acquisitions used to train and test the model and investigate the relationship between their adherence to PI-RADS v2.1 technical standards and model performance in scenarios (*a*) and (*b*).

## Material and Methods

### Study population

A total of 204 patients (median age 66 years [range, 48-83 years]; 204 men) examined at institution one (INST1) in 2012 served as the primary source for model *development*. Patients were initially screened for prostate cancer (PCa) based on high PSA levels and PI-RADS≥ 3 score and PCa diagnosis was confirmed through an MRI-guided biopsy [14]. A total of 30 men recruited at the same institution (INST1) in a different period of time were used to evaluate the *longitudinal performance* of the model (Figure 1).

**Figure 1.**
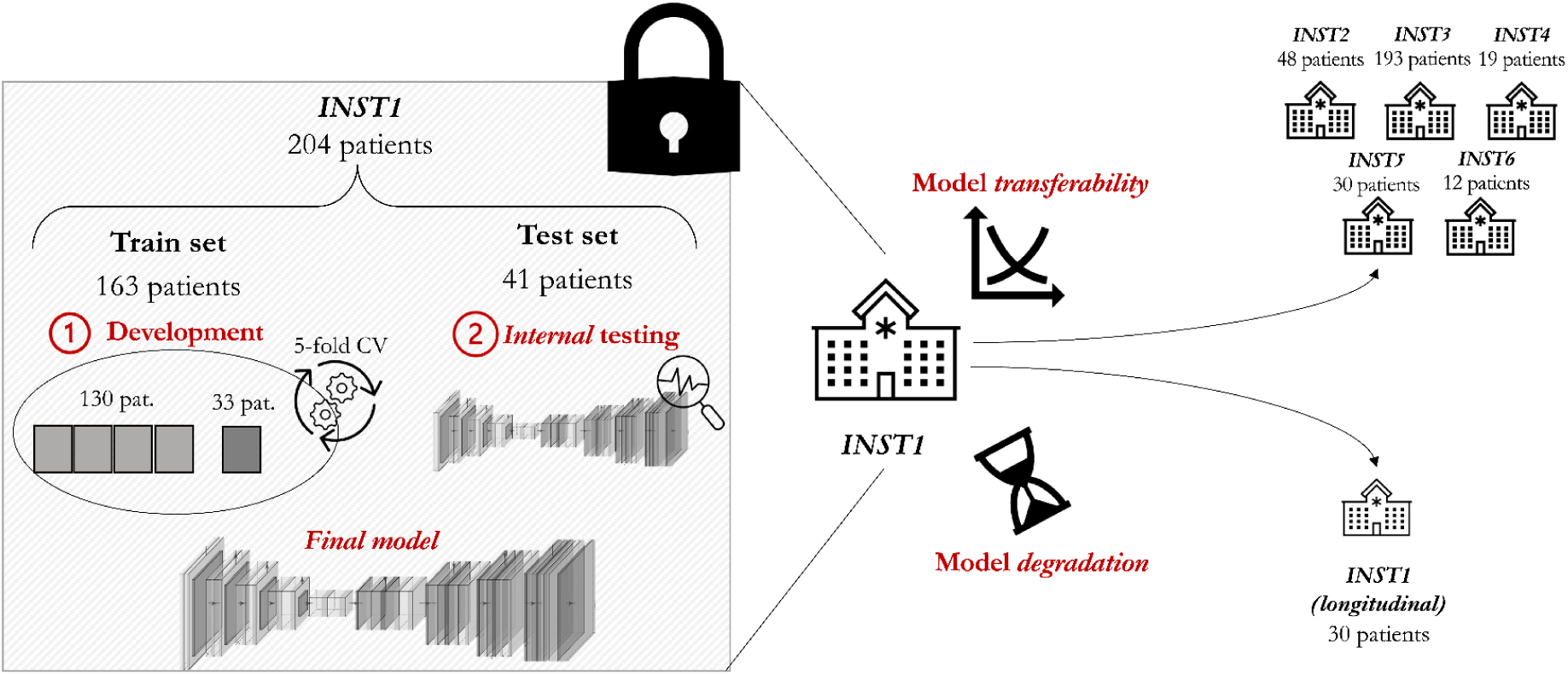
Overview of the approach including model development, transferability and temporal degradation evaluation of the nnU-Net model.

Three groups of patients were treated at institution two (INST2), institution three (INST3) and institution four (INST4) between 2016 and 2019, 2016 and 2020 and between 2003 and 2010, respectively. A total of 48, 139 and 19 patients were included from INST2 (median age 68 years [range, 49-83]; 48 men), INST3 (median age 66 years [range, 35-84 years], 139 men) and INST4 (median age 66 years [range, 40-82 years], 19 men) under the suspicion of PCa based on elevated PSA levels. The three cohorts (INST2, INST3 and INST4) included both PCa biopsy-confirmed cases and negative biopsy cases. Finally, 30 patients from institution 5 (INST5) and 12 patients from institution 6 (INST6) were collected from open-source data [14]. Table 1 A provides a detailed overview of the characteristics of the different cohorts included in the study.

**Table 1:**
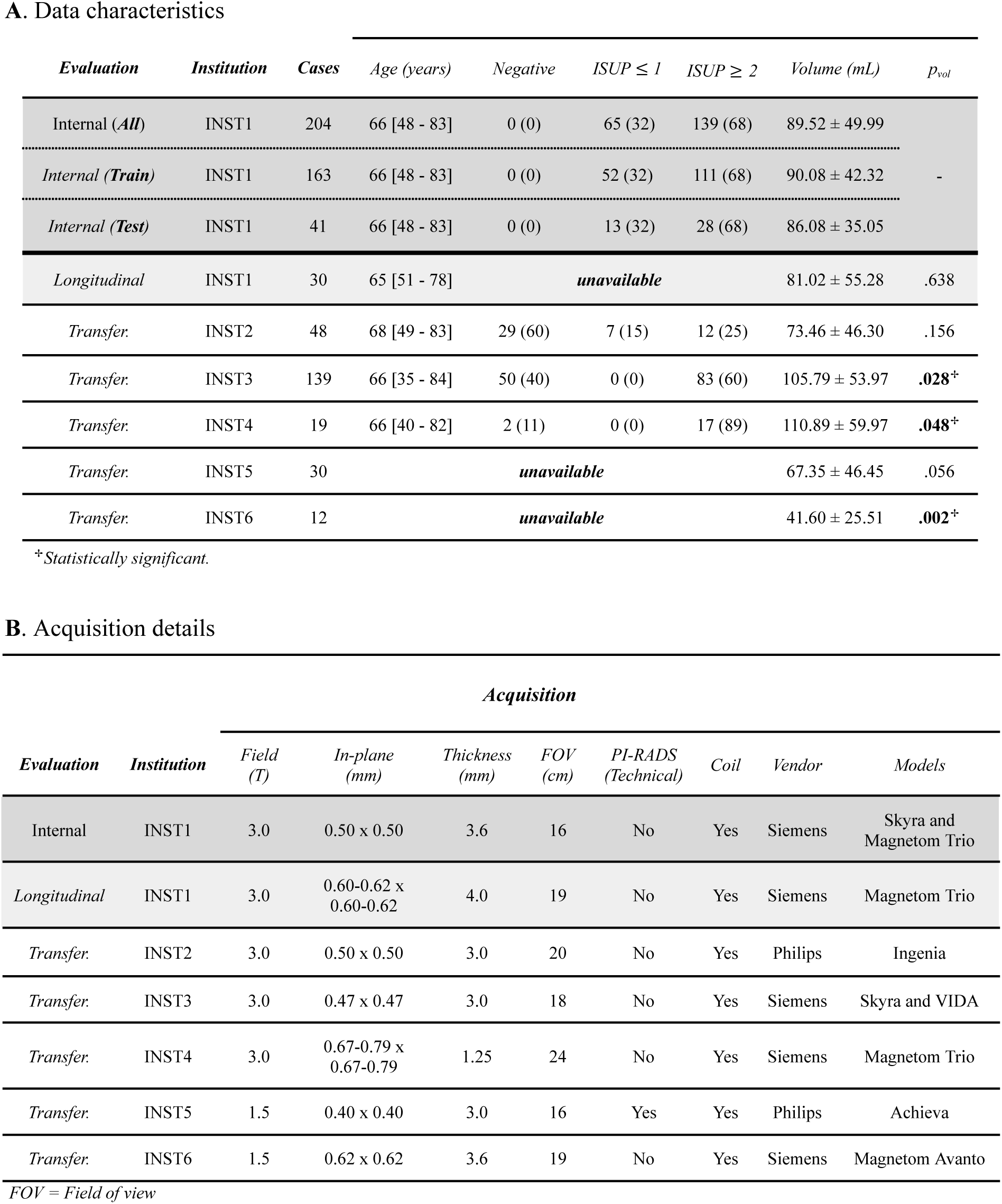
Characteristics of the data used to develop (dark gray) and evaluate the transferability (white) and temporal degradation (light gray). Data is presented as mean (%) or median [range].

A. Data characteristics
| *Evaluation* | *Institution* | *Cases* | *Age (years)* | *Negative* | *ISUP $\leq 1$* | *ISUP $\geq 2$* | *Volume (mL)* | *p<sub>vol</sub>* |
| --- | --- | --- | --- | --- | --- | --- | --- | --- |
| Internal (*All*) | INST1 | 204 | 66 [48 - 83] | 0 (0) | 65 (32) | 139 (68) | 89.52 ± 49.99 | - |
| Internal (*Train*) | INST1 | 163 | 66 [48 - 83] | 0 (0) | 52 (32) | 111 (68) | 90.08 ± 42.32 | |
| Internal (*Test*) | INST1 | 41 | 66 [48 - 83] | 0 (0) | 13 (32) | 28 (68) | 86.08 ± 35.05 | |
| Longitudinal | INST1 | 30 | 65 [51 - 78] | unavailable | | | 81.02 ± 55.28 | .638 |
| Transfer. | INST2 | 48 | 68 [49 - 83] | 29 (60) | 7 (15) | 12 (25) | 73.46 ± 46.30 | .156 |
| Transfer. | INST3 | 139 | 66 [35 - 84] | 50 (40) | 0 (0) | 83 (60) | 105.79 ± 53.97 | **.028<sup>+</sup>** |
| Transfer. | INST4 | 19 | 66 [40 - 82] | 2 (11) | 0 (0) | 17 (89) | 110.89 ± 59.97 | **.048<sup>+</sup>** |
| Transfer. | INST5 | 30 | unavailable | | | | 67.35 ± 46.45 | .056 |
| Transfer. | INST6 | 12 | unavailable | | | | 41.60 ± 25.51 | **.002<sup>+</sup>** |

<sup>+</sup>Statistically significant.

| *Acquisition* |  |  |  |  |  |  |  |  |  |
| --- | --- | --- | --- | --- | --- | --- | --- | --- | --- |
| *Evaluation* | *Institution* | *Field (T)* | *In-plane (mm)* | *Thickness (mm)* | *FOV (cm)* | *PI-RADS (Technical)* | *Coil* | *Vendor* | *Models* |
| Internal | INST1 | 3.0 | 0.50 x 0.50 | 3.6 | 16 | No | Yes | Siemens | Skyra and Magnetom Trio |
| Longitudinal | INST1 | 3.0 | 0.60-0.62 x 0.60-0.62 | 4.0 | 19 | No | Yes | Siemens | Magnetom Trio |
| Transfer. | INST2 | 3.0 | 0.50 x 0.50 | 3.0 | 20 | No | Yes | Philips | Ingenia |
| Transfer. | INST3 | 3.0 | 0.47 x 0.47 | 3.0 | 18 | No | Yes | Siemens | Skyra and VIDA |
| Transfer. | INST4 | 3.0 | 0.67-0.79 x 0.67-0.79 | 1.25 | 24 | No | Yes | Siemens | Magnetom Trio |
| Transfer. | INST5 | 1.5 | 0.40 x 0.40 | 3.0 | 16 | Yes | Yes | Philips | Achieva |
| Transfer. | INST6 | 1.5 | 0.62 x 0.62 | 3.6 | 19 | No | Yes | Siemens | Magnetom Avanto |

FOV = Field of view

### Image acquisition and manual annotations

T2-weighted (T2w) axial sequences were considered as the main imaging data source. The nature of all the imaging data is retrospective. Sequences used for model *development* (INST1) were acquired with a 3.0-Tesla (T) Siemens scanner with an in-plane resolution of 0.5 mm x 0.5 mm, 3.6 mm slice thickness and with a surface coil. The acquisitions were *not* consistent with PI-RADS v.2.1 recommendations, as presented in Table 2. Examinations from other institutions presented an heterogeneous technical acquisition: INST2 were acquired with an in-plane resolution of 0.5 mm x 0.5 mm, 3.0 mm slice thickness whilst INST4 sequences were acquired with an in-plane resolution of 0.67 -0.79 mm x 0.67-0.79 mm and 1.25 mm slice thickness, which was *not* consistent with the technical recommendations. Table 1B and Table 2 depict an in-depth summary of the technical acquisition characteristics and vendors from different data sources included in the study.

**Table 2:** PI-RADS v2.1 technical adherence of the data used to develop (dark gray) and evaluate the transferability (white) and temporal degradation (light gray) of the model.

| *PI-RADS technical standards adherence* |  |  |  |  |  |  |
| --- | --- | --- | --- | --- | --- | --- |
| *Evaluation* | *Institution* | *Phase*<br>$\leq 0.7 \text{ mm}$ | *Frequency*<br>$\leq 0.4 \text{ mm}$ | *Thickness*<br>$= 3 \text{ mm}$ | $12\text{cm} \leq \text{FOV} \leq 20 \text{ cm}$ | *Total* |
| Internal | INST1 | Yes | No | No | Yes | 2/4 |
| Longitudinal | INST1 | Yes | No | No | Yes | 2/4 |
| Transfer. | INST2 | Yes | No | Yes | Yes | 3/4 |
| Transfer. | INST3 | Yes | No | Yes | Yes | 3/4 |
| Transfer. | INST4 | No | No | No | No | 0/4 |
| Transfer. | INST5 | Yes | Yes | Yes | Yes | 4/4 |
| Transfer. | INST6 | Yes | No | No | Yes | 2/4 |

*FOV = Field of view*

Included manual annotations of the whole gland were performed by two radiologists in-training and two experienced board-certified radiologists for INST1 [15], by a radiologist in-training (INST2), two-board certified radiologists (INST3), by one experienced radiologist (> 5 years, INST4 and INST6) and by a panel of 5 experienced radiologists (>5 years, INST1 longitudinal and INST5), respectively. In the case of INST1, INST2, INST3 and INST4 segmentations were obtained using ITKSnap v3.8 whilst INST5 and INST6 used MeVisLab and 3DSlicer. The Regional Committee for Medical and Health Research Ethics (REC Central Norway) approved the use of the INST2 dataset (2019/272). All the patients signed informed consent prior to the collection of the data.

### Pre-processing and data splitting

As depicted in Figure 1, INST1 data is used to *train* and *develop* the models. We use one of the most used *open-source* and *single-institution* datasets in the literature to develop prostate WG segmentation models [10, 14, 16]. We split INST1 data in 80/20% to obtain the train set and *internal* test set, resulting in 163 and 41 patients, respectively. The splitting is performed ensuring no cross-contamination. Different centers employ different acquisition protocols, leading to large inter-site differences. To alleviate those differences, the pixel intensity range is normalized followed by a center cropping, axis re-ordering and a resampling of the sequences. All the steps are part of the pre-processing pipeline of nn-UNet [15].

### Segmentation model

We train a standard nn-UNet [15] based on its wide adoption and results in prostate segmentation challenges. Training is performed with the default configuration of the network, as exploration of architecture modifications is considered out of the scope of work: a loss that combines dice loss and cross-entropy, SGD optimizer, trained for 1000 epochs and with an automatic selection of data augmentation techniques applied *on the fly* [15]. The training is performed with a 5-fold cross-validation (CV), where the final segmentation model used in the study is the result of ensembling the 5 models obtained from each iteration in the CV. The model is implemented using Tensorflow and Python language. We train and evaluate the model on an NVIDIA A100-80G GPU.

### Evaluation

#### Prostate segmentation

We evaluate the nn-UNet model trained with data from INST1 in terms of *transferability* and *longitudinal degradation* with data from other institutions (INST2, INST3, INST4, INST5 and INST6), and data from the same institution acquired in a different period of time and with different technical characteristics (Figure 2). Evaluations are carried out using the model *as given* (out-of-the-box), under the **assumption** the receiving institutions do not have the possibility nor resources to obtain manual annotations or to perform any calibration (*re-training*) of the model, as depicted in Figure 1.

**Figure 2.**
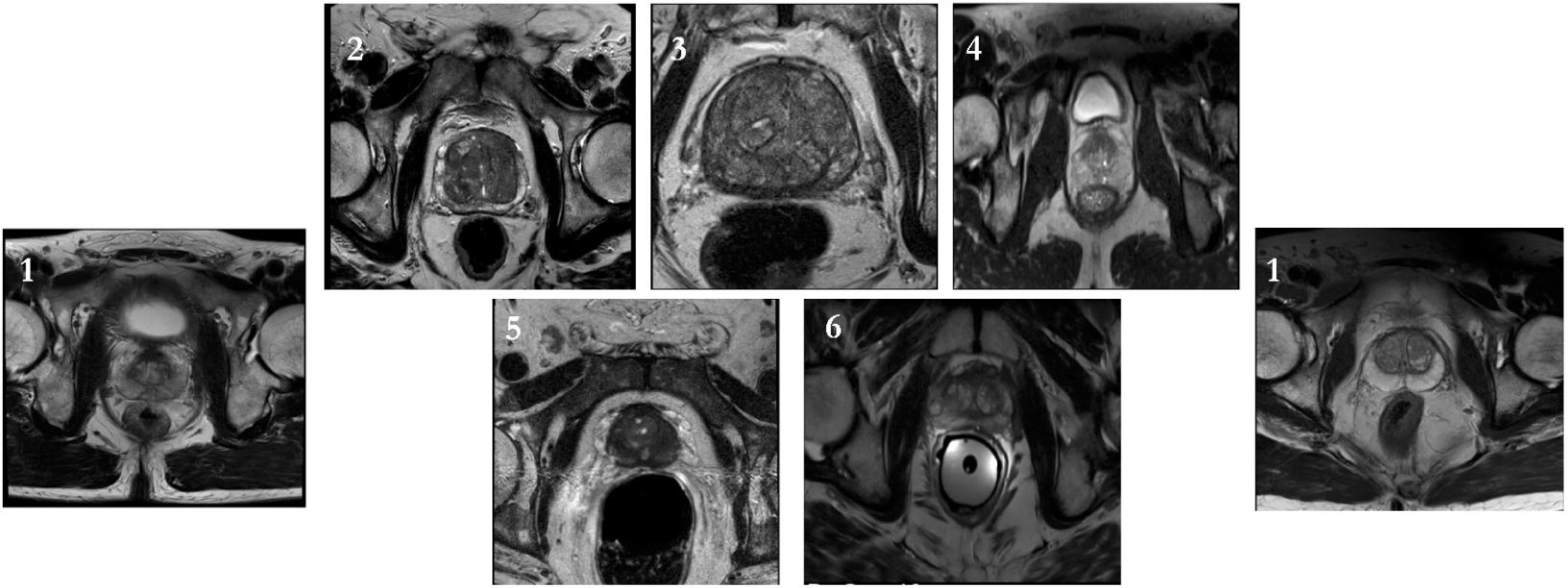
Examples of T2w axial prostate MRI scans from the different institutions depicting the inter-institutional qualitative heterogeneity: *development* data followed by evaluation. Number indicates the institution identifier (i.e. INST1).

The results are presented with 100 bootstrap replicates without repetition in the form of mean ± standard deviation (SD) [17]. To focus on clinically relevant measures, we present results at the *patient level* for different prostate regions of interest: *WG*, *base*, *mid-gland* and *apex*. Each region is assumed to contain ⅓ of the total number of MRI slices *containing the prostate* in a given patient sequence [9]. We quantify the quality of the segmentation through Dice Score Coefficient (DSC, %), Hausdorff distance (HD, mm), Absolute Boundary Distance (ABD, mm) and Relative Volume Difference (RVD, %) [9].

#### Statistical analysis

We present quantitative results for all the metrics and prostate regions included in study. Specifically, we compare the results for DSC and RVD at the whole patient level through a permutation test with *n* = 1000 repetitions, to avoid assumptions of the underlying distributions. Pearson correlation coefficient is used to evaluate the linear relationship between predicted volumes and ground truth. Baseline was defined as the results originating from the *internal test* performance of INST1 dataset. We consider a *P* value less than .05 to be statistically significant.

## Results

### Differences in acquisition

The different *external* institutions presented high variability in acquisition parameters and adherence to PI-RADS v2.1 technical standards. As depicted in Table 1B, 60% of the T2w axial sequences used for the *transferability study* were acquired with Siemens MRI scanners (INST3, INST4 and INST6), whilst the rest of institutions (40%) performed the acquisitions with Philips scanners (INST2 and INST5). Despite using the same vendor, the *longitudinal analysis* accounted for changes in models, where the data used to train the model was acquired with both Siemens Skyra and Siemens Magnetom Trio and the *longitudinal test data* was acquired with Siemens Magnetom Trio, exclusively. Specifically, a 75% of the Siemens and a 50% of the Philips acquisitions were performed with a 3.0 T field strength (INST1, INST2, INST3 and INST4) whilst a 25% and 50% were performed with 1.5 T field strength, respectively (INST5 and INST6). Whilst all of the different institutions used different models, Siemens Magnetom Trio was the most common one (INST1 and INST4).

Remarkably, the different institutions presented a different degree of adherence to PI-RADSv2.1 technical standards. In particular, 50% (INST2, INST3 and INST5) performed the acquisitions with the recommended 3.0 mm slice thickness whilst 50% (INST1, INST4 and INST6) did not follow the recommendations. All of the institutions used an in-plane phase dimension ≤ 0.7 mm except for INST4 (17%) whilst only one institution (INST5) presented adherence to the recommended in-plane frequency≤0.4 mm (INST5). All the institutions adhered to the field of view (FOV) 12-20 cm recommendations by PI-RADSv2.1 except for INST4 (Table 2). As depicted in Table 1A, the WG volume distributions of the populations included in the study presented significantly larger (INST3 and INST4) and significantly lower volumes (INST6) when compared to that of the data used to train the algorithm.

### Transferability and MRI scanner characteristics

Table 3 presents the main results for the model *transferability* and *longitudinal* performance, respectively, whilst Figure 3 presents qualitative results for the different institutions and studies. Results for all the prostate regions are presented in Appendix A. In the *transferability study*, the model presented a significantly lower WG DSC for all institutions when compared to the baseline (89.78 ± 5.420% [standard error] INST2 *P<* .001, 75.75±14.56% INST3 *P<* .001, 74.83±15.04% INST4 *P<*.001, 80.57±11.87% INST5 *P<*.001 and 77.94±12.71% INST6 *P<*.001) and significantly lower RVD for five different institutions (-2.588 ± 12.40% INST2 *P=* .003, -27.74±21.46% INST3 *P<*.001, -22.20±19.79% INST5 *P<*.001, -18.80±19.09% INST6 *P<*.001).

**Figure 3.**
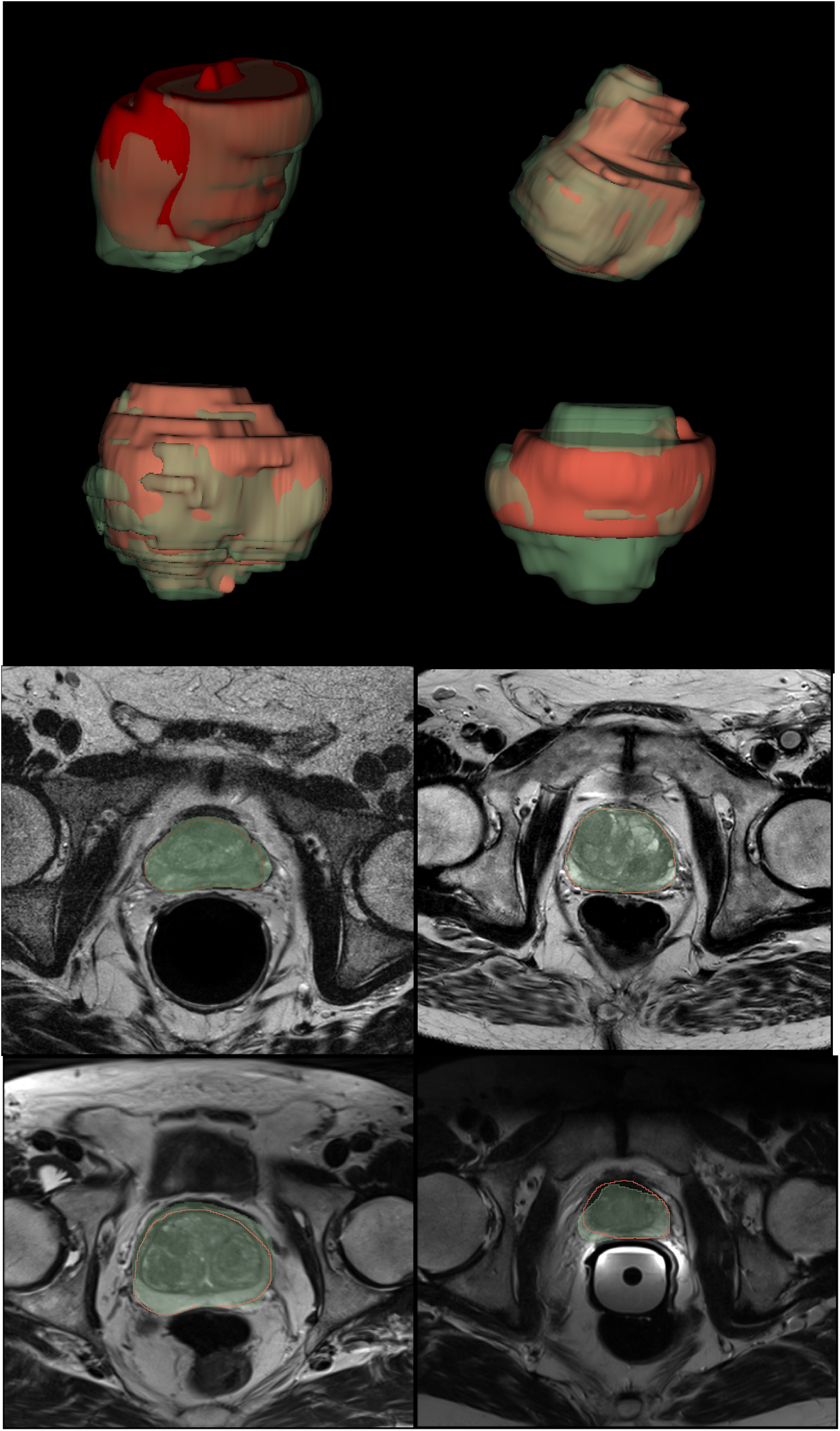
Qualitative segmentation results. Upper row depicts the 3D rendering of the predicted prostate segmentation blended with the expert annotation, in green and red, respectively. Bottom rows show the slice-level predictions. From left to right: INST5, INST2, INST1 (longitudinal) and INST6.

**Table 3:** Transferability and temporal degradation WG segmentation results with *n* = 100 bootstrap replicates without repetition. Results are presented as mean ± SD.

| *Institution* | *Region* | *Metrics* | | | | $P_{DSC}^{**}$ | $P_{RVD}^{**}$ |
| --- | --- | --- | --- | --- | --- | --- | --- |
|  |  | *DSC (%)* ↑ | *HD (mm)* ↓ | *ABD (mm)* ↓ | *RVD (%)* ↓ |  |  |
| *INST1*<br>(*Test*) | **WG** | 99.46 ± 0.125 | 2.16 ± 1.30 | 0.030 ± 0.006 | -0.006 ± 0.127 | - |  |
|  | Base | 99.40 ± 0.163 | 1.42 ± 0.50 | 0.015 ± 0.004 | -0.065 ± 0.182 |  |  |
|  | Mid | 99.54 ± 0.113 | 1.83 ± 1.26 | 0.017 ± 0.003 | 0.023 ± 0.130 |  |  |
|  | Apex | 99.17 ± 0.375 | 1.50 ± 0.75 | 0.014 ± 0.005 | -0.038 ± 0.495 |  |  |
| *INST1*<br>(*Longitudinal*) | **WG** | 91.24 ± 3.320 | 8.02 ± 4.43 | 0.68 ± 0.26 | 8.10 ± 8.16 | < .001<sup>+</sup> | < .001<sup>+</sup> |
|  | Base | - | - | - | - | - | - |
|  | Mid | 93.83 ± 4.470 | 6.77 ± 2.71 | 0.285 ± 0.145 | 4.48 ± 10.57 |  |  |
|  | Apex | - | - | - | - |  |  |
| *INST2*<br>(*Transfer.*) | **WG** | 89.78 ± 5.420 | 10.31 ± 11.67 | 1.130 ± 3.161 | -2.588 ± 12.40 | < .001<sup>+</sup> | .003<sup>+</sup> |
| *INST3*<br>(*Transfer.*) | **WG** | 75.75 ± 14.56 | 15.52 ± 9.47 | 1.490 ± 0.693 | -27.74 ± 21.46 | < .001<sup>+</sup> | < .001<sup>+</sup> |
| *INST4*<br>(*Transfer.*) | **WG** | 74.83 ± 15.04 | 17.32 ± 6.85 | 3.20 ± 1.02 | 1.187 ± 28.34 | < .001<sup>+</sup> | 0.674 |
| *INST5*<br>(*Transfer.*) | **WG** | 80.57 ± 11.87 | 16.52 ± 21.70 | 1.16 ± 0.44 | -22.20 ± 19.79 | < .001<sup>+</sup> | < .001<sup>+</sup> |
| *INST6*<br>(*Transfer.*) | **WG** | 77.94 ± 12.71 | 10.28 ± 6.42 | 1.22 ± 0.41 | -18.80 ± 19.09 | < .001<sup>+</sup> | < .001<sup>+</sup> |

*DSC* = Dice score coefficient, *HD* = Hausforff distance, *ABD* = Absolute boundary distance and *RVD* = relative volume difference
<sup>\*\*</sup>Reference is *INST1* (internal test, first row).
<sup>+</sup>Statistically significant.

As depicted in Figures 4 and 5, when assessing the effect of the acquisition characteristics and scanners and their impact in the *transferability* of the model, a significantly lower WG DSC and RVD can be observed for Siemens when compared to Philips (86.24±9.67 Philips and 75.81±14.55 Siemens, *P<*.001) and (-10.13±18.46% Philips and -23.88±24.08% Siemens, *P<*.001). Further, differences were observed in the different vendor models, with a particularly significantly lower DSC for the Siemens trio scanner when compared to the baseline (74.83±15.04% *P<*.001). In particular, when analyzing the field strength, no differences were observed for scanners using a 3.0T or 1.5T field strength (DSC: 78.94±14.41% and 79.82±12.32%, *P=*.712; RVD: -19.21±23.99% and -21.23±19.89%, *P=*.609).

**Figure 4.**
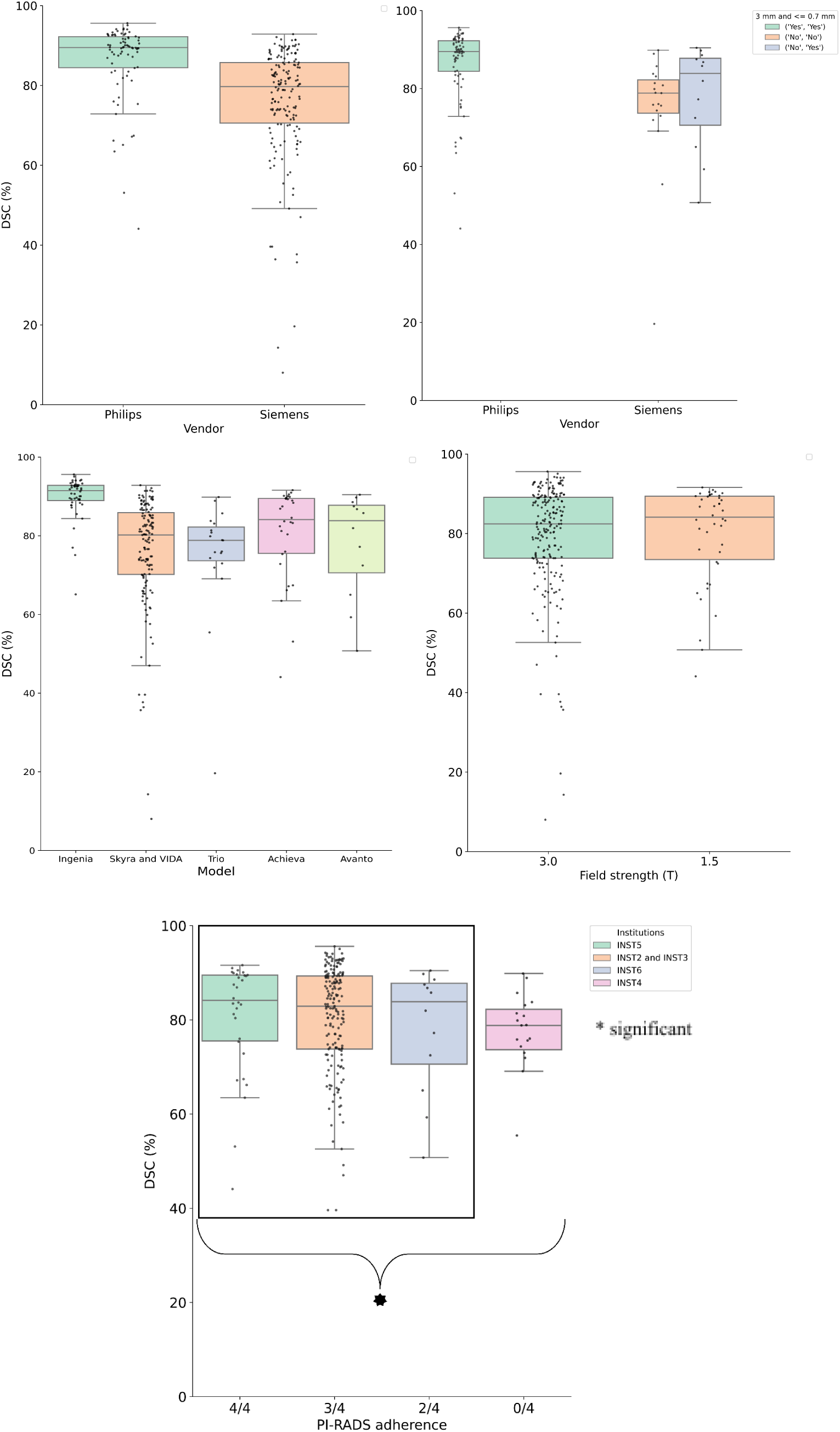
Dice score (%) of the whole gland by scanner characteristics: field strength (T), Vendor, scanner model and PI-RADS v2.1 adherence.

**Figure 5.**
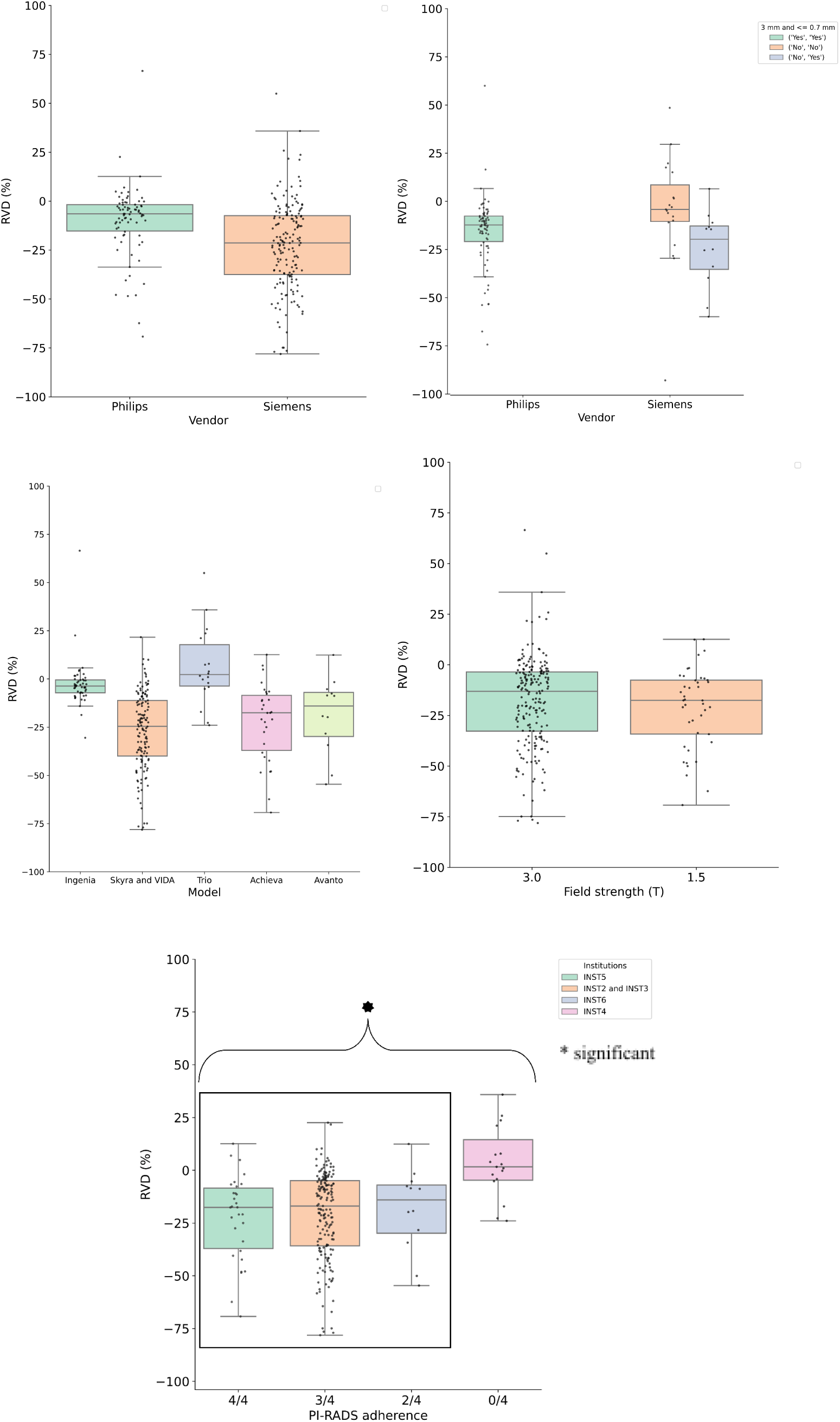
Relative volume difference (%) of the whole gland by scanner characteristics: field strength (T), Vendor, scanner model and PI-RADS v2.1 adherence.

### PI-RADSv2.1 technical adherence

Whilst Siemens and Philips scanners present significant differences in the *transferability* of the model, we observe that those differences can be explained by the degree of adherence to PI-RADSv.2.1 technical standards of the different institutions. To be more specific, as depicted in Figure 4 and 5, the different MRI scanner model protocols present different degrees of adherence to the technical recommendations of PI-RADS. Scanners presenting adherence to slice thickness and in-plane phase recommendations present a significantly higher DSC and RVD when compared to those not adhering to them (DSC: 86.24±9.67 % and 74.83±15.45 % *P<*.001; RVD: -6.50±18.46 % and 1.64±29.12 % *P=* .003). The same trend can be observed when compared those following, exclusively, slice thickness recommendations when compared to those presenting a slice thickness≥3mm (DSC: 86.24±9.67 % and 77.94±13.27 % *P<* .001; RVD: -6.50±18.46 % and -14.03±19.94 % *P<*.001). Further, a significant difference is also observed when comparing those scanners following only one technical recommendation (thickness≤3 mm, frequency≤0.4 mm or phase≤0.7 mm) with those not following any (DSC: 77.94±13.27 % and 74.83±15.45 % *P<* .001; RVD: -14.03±19.94 % 1.64±29.12 % *P* <.001). The effect of PI-RADS v2.1 technical recommendations adherence can also be observed in Appendix A, where base and apex are commonly disregarded and segmentation predictions are only obtained for the mid region of the WG.

### Prostate volume distribution and the effect on transferability

Using the segmentation masks, the prostate whole gland volume could be estimated for each patient. As shown in Figure 6, we observe significant differences in the spearman correlation coefficient between the volume obtained from the predicted WG masks for each institution and the ground truth volume, with the lowest value observed for INST4 (0.56, *P*<.001) followed by INST5, INST6, INST3 and INST2, respectively (0.73 *P*<.001; 0.88 *P*<.001; 0.88 *P*<.001; 0.95 *P*<.001). We also observe a good spearman correlation for the *longitudinal analysis* (0.97, *P*<.001).

**Figure 6.**
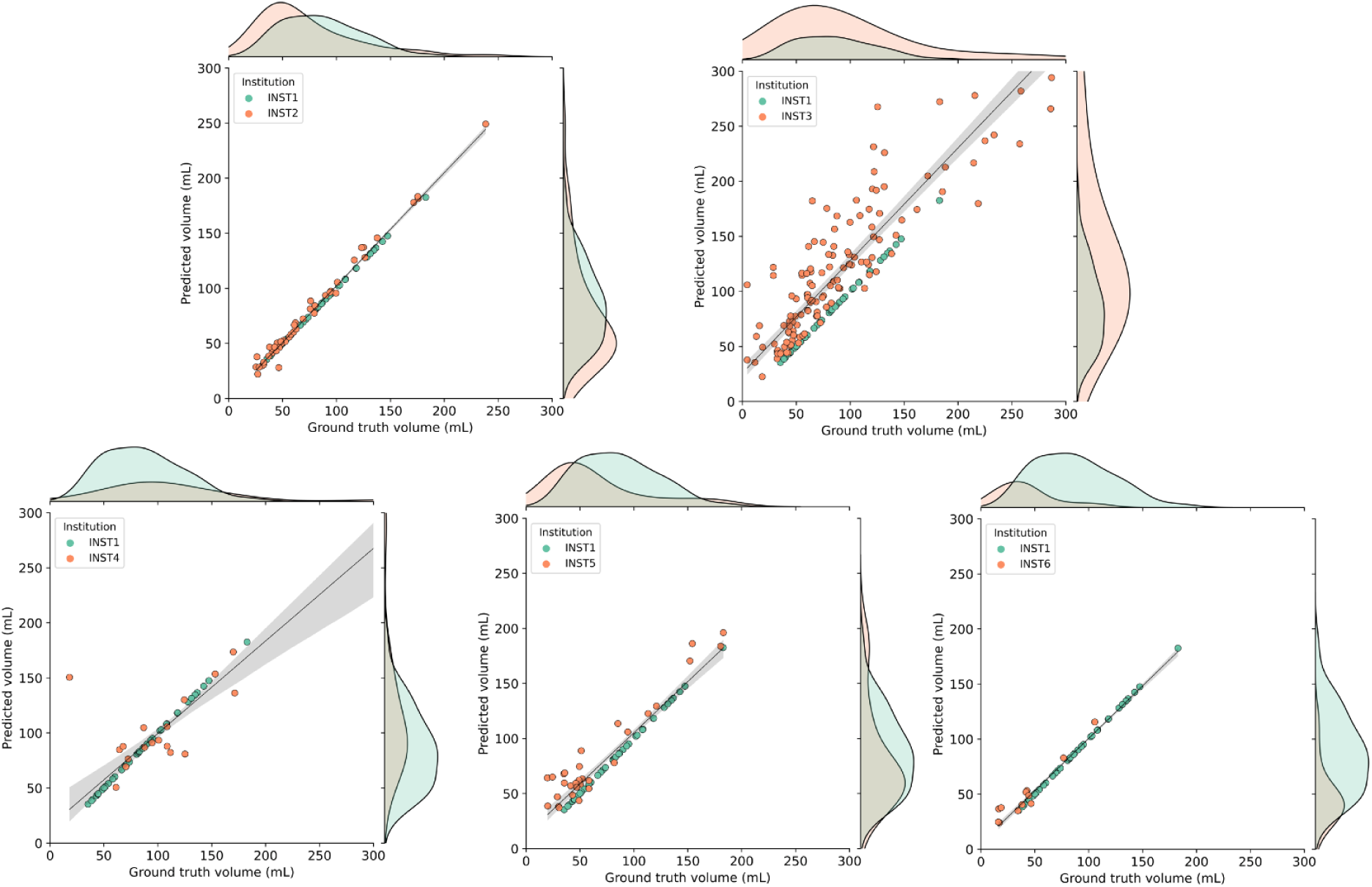
Prostate whole gland volume prediction (mL) per patient compared to the ground truth for the transferability study, including volume distribution plots. Regressions are displayed with their *n* = 1000 bootstrap estimated 95% confidence interval.

### Model continuous monitoring

We investigated the effect on the performance of an already deployed model in the event of a modification of technical acquisition parameters or scanner model. Table 2 (longitudinal) depicts the results obtained for the *longitudinal performance*. In particular, we observe a significantly lower average DSC when compared to the baseline: 99.46±0.12% and 91.24±3.32% (*P*<.001) and a significantly lower RVD:-0.006±0.127% and 8.10±8.16% (*P*<.001). Further, we can observe the challenges faced by the algorithm in the base and the apex regions of the prostate, where segmentations are not obtained. Qualitative results for the *longitudinal segmentation performance* are shown in Figure 3.

## Discussion

In radiology and oncological imaging, scenarios reflecting the intended AI use-case and carefully evaluating clinical applicability in *simulated deployment* are seldomly accounted for. Traditionally, the development and deployment of an AI tool is already considered a success and post-deployment (*longitudinal*) assessments of the performance are not carried out, in spite of the dynamic nature of clinical environments [22, 23, 24, 25]. For that purpose, we train through a 5-cross validation an AI prostate WG segmentation algorithm using data from one institution commonly used by many existing studies as their main data development source [9]. Following, we focus on analyzing the *transferability* and *longitudinal performance* of the developed algorithm under different imaging acquisition characteristics. We compare the technical acquisition characteristics against standardized recommendations by PI-RADSv2.1 and under realistic conditions, where annotated data is commonly not available or limited at the receiving institution and consequently, possibility of algorithm re-calibration is not available [22, 24].

Our results depict a significant decrease in DSC performance when the developed algorithm is *transferred* outside of the institution where it was developed (DSC:99.46±0.125% [standard error] INST1 and 89.78±5.420% [standard error] INST2,*P<*.001). Further, a significant volume estimation performance decrease is observed under the same conditions (RVD:-0.006±0.127% INST1 and -2.588±12.40% INST2,*P=*.003). Of particular importance is the latter, where prostate cancer risk markers directly depend on the estimation and accuracy of the prostate whole gland volume [4].

We observe similar trends as in the *transferability* study when assessing the *post-deployment performance* of the algorithm at the same institution where it was developed under different acquisition and scanner characteristics. Specifically, there is a significant decrease in DSC and RVD performance (DSC:99.46±0.125% and 91.24±3.32%, *P*<.001; RVD:-0.006±0.127% and 8.10±8.16% *P*<.001). More importantly, we observe that both when *transferred to external institutions* or tested under *acquisition shifts at the same institution* the algorithm considerably underperforms in the apex and base regions of the prostate, presumably due to the differences in slice thickness and in-plane resolutions of the scans that may increase the partial volume effect, making the prostate capsule more obscured and harder to distinguish for the algorithm.

When investigating the association between PI-RADSv2.1 technical adherence of the different institutions and the previously reported differences in *transferability* and *post-deployment* study, we observe a positive impact in institutions that present a higher degree of adherence. In particular, the higher the degree of adherence measured by a 3.0 mm slice thickness, in-plane phase dimension≤0.7 mm and in-plane frequency≤0.4 mm, the higher the DSC and RVD performance: (DSC:86.24±9.67% and 77.94±13.27%,*P<*.001; RVD: -6.50±18.46 % and -14.03±19.94%,*P<*.001 *full adherence and only slice thickness*) and (DSC:77.94±13.27% and 74.83±15.45%,*P<*.001; RVD:-14.03±19.94% 1.64±29.12 %,*P*<.001 *only slice thickness vs no adherence*).

Our study had several limitations, including the limited case-scenario of one training dataset and architecture, which could limit the generalizability of the results. Although the dataset used to train the model is widely adopted, it was acquired in 2012, which limits its technical adherence and might negatively impact the model performance. Nevertheless, based on previous reports, there are reasons to think similar results would be obtained for different datasets and algorithms. Other limitations included the amount of data in the post-deployment scenario, where ideally, the simulation could be performed with more data and in a prospective way. Finally, missing descriptive data for INST5 and INST6 can also limit the interpretability and control of potential cofounders in the study.

In conclusion, the degree of adherence to PI-RADSv2.1 technical acquisition recommendations may affect the performance of prostate WG segmentation *inter-institutional* transferability and *post-deployment* at a given institution. Our results highlight the importance of continuous model monitoring and standardized technical acquisition protocols in agreement with PI-RADSv2.1 guidelines recommendations for those institutions adopting AI-assisted prostate segmentation.

## Data Availability

All data produced in the present study are available upon reasonable request to the authors.

## APPENDIX A

**Table 3:** Region-based transferability and temporal degradation segmentation results with *n* = 100 bootstrap replicates without repetition. Results are presented as mean ± SD.

| Institution | Region | Metrics | | | | $P_{DSC}^{**}$ | $P_{RVD}^{**}$ |
| --- | --- | --- | --- | --- | --- | --- | --- |
| | | DSC (%) ↑ | HD (mm) ↓ | ABD (mm) ↓ | RVD (%) ↓ | | |
| INST1<br>(Test) | WG | 99.46 ± 0.125 | 2.16 ± 1.30 | 0.030 ± 0.006 | -0.006 ± 0.127 | - | |
| | Base | 99.40 ± 0.163 | 1.42 ± 0.50 | 0.015 ± 0.004 | -0.065 ± 0.182 | | |
| | Mid | 99.54 ± 0.113 | 1.83 ± 1.26 | 0.017 ± 0.003 | 0.023 ± 0.130 | | |
| | Apex | 99.17 ± 0.375 | 1.50 ± 0.75 | 0.014 ± 0.005 | -0.038 ± 0.495 | | |
| INST1<br>(Longitudinal) | WG | 91.24 ± 3.32 | 8.02 ± 4.43 | 0.68 ± 0.26 | 8.10 ± 8.16 | < .001<sup>+</sup> | < .001<sup>+</sup> |
|  | Base | - | - | - | - | - | - |
| | Mid | 93.83 ± 4.47 | 6.77 ± 2.71 | 0.285 ± 0.145 | 4.48 ± 10.57 | | |
|  | Apex | - | - | - | - |  |  |
| INST2<br>(Transfer.) | WG | 89.78 ± 5.42 | 10.31 ± 11.67 | 1.130 ± 3.161 | -2.588 ± 12.40 | < .001<sup>+</sup> | .003<sup>+</sup> |
|  | Base | - | - | - | - | - | - |
| | Mid | 91.05 ± 6.39 | 10.20 ± 11.72 | 0.863 ± 3.192 | 1.761 ± 15.95 | | |
|  | Apex | - | - | - | - |  |  |
| INST3<br>(Transfer.) | WG | 75.75 ± 14.56 | 15.52 ± 9.47 | 1.490 ± 0.693 | -27.74 ± 21.46 | < .001<sup>+</sup> | < .001<sup>+</sup> |
|  | Base | - | - | - | - | - | - |
| | Mid | 79.27 ± 17.74 | - | - | -23.76 ± 25.37 | | |
|  | Apex | - | - | - | - |  |  |
| INST4<br>(Transfer.) | WG | 74.83 ± 15.04 | 17.32 ± 6.85 | 3.20 ± 1.02 | 1.187 ± 28.34 | < .001<sup>+</sup> | 0.674 |
|  | Apex | - | - | - | - | - | - |
| | Mid | 78.79 ± 15.17 | 7.433 ± 2.448 | 2.44 ± 1.04 | -4.59 ± 28.21 | | |
|  | Base | - | - | - | - |  |  |
| INST5<br>(Transfer.) | WG | 80.57 ± 11.87 | 16.52 ± 21.70 | 1.16 ± 0.44 | -22.20 ± 19.79 | < .001<sup>+</sup> | < .001<sup>+</sup> |
|  | Apex | - | - | - | - | - | - |
| | Mid | 83.90 ± 13.09 | 16.41 ± 22.78 | 0.703 ± 0.422 | -17.74 ± 20.24 | | |
|  | Base | - | - | - | - |  |  |
| INST6<br>(Transfer.) | WG | 77.94 ± 12.71 | 10.28 ± 6.42 | 1.22 ± 0.41 | -18.80 ± 19.09 | < .001<sup>+</sup> | < .001<sup>+</sup> |
|  | Apex | - | - | - | - | - | - |
| | Mid | 84.93 ± 10.65 | 10.26 ± 6.44 | 0.65 ± 0.38 | -6.35 ± 16.53 | | |
|  | Base | - | - | - | - |  |  |

<sup>\*\*</sup>Reference is INST1 (internal test, first row). <sup>+</sup>Statistically significant.

## References

1. Wu C, Montagne S, Hamzaoui D, Ayache N, Delingette H, Renard-Penna R. Automatic segmentation of prostate zonal anatomy on MRI: a systematic review of the literature. Insights into Imaging. 2022 Dec;13(1):1–7. doi.org/10.1186/s13244-022-01340-2

2. Kwee TC, Kwee RM. Workload of diagnostic radiologists in the foreseeable future based on recent scientific advances: growth expectations and role of artificial intelligence. Insights into imaging. 2021 Dec;12(1):1–2. doi.org/10.1186/s13244-021-01031-4

3. Lamy PJ, Allory Y, Gauchez AS, Asselain B, Beuzeboc P, De Cremoux P, Fontugne J, Georges A, Hennequin C, Lehmann-Che J, Massard C. Prognostic biomarkers used for localised prostate cancer management: a systematic review. European Urology Focus. 2018 Dec 1;4(6):790–803. doi.org/10.1016/j.euf.2017.02.017

4. Noguchi M, Stamey TA, McNeal JE, Yemoto CM. Relationship between systematic biopsies and histological features of 222 radical prostatectomy specimens: lack of prediction of tumor significance for men with nonpalpable prostate cancer. The Journal of urology. 2001 Jul;166(1):104–10. PMID: 11435833

5. Fernandez-Quilez A, Nordström T, Jäderling F, Kjosavik SR, Eklund M. Prostate Age Gap (PAG): An MRI surrogate marker of aging for prostate cancer detection. arXiv preprint arXiv:2308.05344. 2023 Aug 10.

6. Becker AS, Chaitanya K, Schawkat K, Muehlematter UJ, Hötker AM, Konukoglu E, Donati OF. Variability of manual segmentation of the prostate in axial T2-weighted MRI: A multi-reader study. European journal of radiology. 2019 Dec 1;121:108716. doi.org/10.1016/j.ejrad.2019.108716

7. Fernandez-Quilez A, Larsen SV, Goodwin M, Gulsrud TO, Kjosavik SR, Oppedal K. Improving prostate whole gland segmentation in t2-weighted mri with synthetically generated data. In2021 IEEE 18th International Symposium on Biomedical Imaging (ISBI) 2021 Apr 13 (pp. 1915–1919). IEEE. doi.org/10.1109/ISBI48211.2021.9433793

8. Isensee F, Jaeger PF, Kohl SA, Petersen J, Maier-Hein KH. nnU-Net: a self-configuring method for deep learning-based biomedical image segmentation. Nature methods. 2021 Feb;18(2):203–11. doi.org/10.1038/s41592-020-01008-z

9. Litjens G, Toth R, Van De Ven W, Hoeks C, Kerkstra S, van Ginneken B, Vincent G, Guillard G, Birbeck N, Zhang J, Strand R. Evaluation of prostate segmentation algorithms for MRI: the PROMISE12 challenge. Medical image analysis. 2014 Feb 1;18(2):359–73. doi.org/10.1016/j.media.2013.12.002

10. Lindeijer TM, Ytredal TM, Eftestøl T, Nordström T, Jäderling F, Eklund M, Fernandez-Quilez A. Leveraging multi-view data without annotations: a contrastive approach. arXiv preprint arXiv:2308.06477. 2023 Aug 12.

11. Giganti F, Kirkham A, Kasivisvanathan V, Papoutsaki MV, Punwani S, Emberton M, Moore CM, Allen C. Understanding PI-QUAL for prostate MRI quality: a practical primer for radiologists. Insights into Imaging. 2021 Dec;12(1):1–9.

12. Rauschecker AM, Gleason TJ, Nedelec P, Duong MT, Weiss DA, Calabrese E, Colby JB, Sugrue LP, Rudie JD, Hess CP. Interinstitutional portability of a deep learning brain MRI lesion segmentation algorithm. Radiology: Artificial Intelligence. 2021 Nov 10;4(1):e200152.

13. Vela D, Sharp A, Zhang R, Nguyen T, Hoang A, Pianykh OS. Temporal quality degradation in AI models. Scientific Reports. 2022 Jul 8;12(1):11654.

14. Litjens G, Debats O, Barentsz J, Karssemeijer N, Huisman H. Computer-aided detection of prostate cancer in MRI. IEEE transactions on medical imaging. 2014 Jan 30;33(5):1083–92. doi.org/10.1109/TMI.2014.2303821

15. Isensee F, Jaeger PF, Kohl SA, Petersen J, Maier-Hein KH. nnU-Net: a self-configuring method for deep learning-based biomedical image segmentation. Nature methods. 2021 Feb;18(2):203–11.

16. Cuocolo R, Comelli A, Stefano A, Benfante V, Dahiya N, Stanzione A, Castaldo A, De Lucia DR, Yezzi A, Imbriaco M. Deep learning whole-gland and zonal prostate segmentation on a public MRI dataset. Journal of Magnetic Resonance Imaging. 2021 Aug;54(2):452–9. doi.org/10.1002/jmri.27585

17. Fernandez-Quilez A, Eftestøl T, Kjosavik SR, Goodwin M, Oppedal K. Contrasting Axial T2W MRI for Prostate Cancer Triage: A Self-Supervised Learning Approach. In 2022 IEEE 19th International Symposium on Biomedical Imaging (ISBI) 2022 Mar 28 (pp. 1–5). IEEE. doi.org/10.1109/ISBI52829.2022.9761573.

18. Srivastava N, Hinton G, Krizhevsky A, Sutskever I, Salakhutdinov R. Dropout: a simple way to prevent neural networks from overfitting. The journal of machine learning research. 2014 Jan 1;15(1):1929–58.

19. Eklund M, Jäderling F, Discacciati A, Bergman M, Annerstedt M, Aly M, Glaessgen A, Carlsson S, Grönberg H, Nordström T. MRI-targeted or standard biopsy in prostate cancer screening. New England journal of medicine. 2021 Sep 2;385(10):908–20.

20. Li H, Lee CH, Chia D, Lin Z, Huang W, Tan CH. Machine learning in prostate MRI for prostate cancer: current status and future opportunities. Diagnostics. 2022 Jan 24;12(2):289. doi.org/10.3390/diagnostics12020289

21. Hosseinzadeh M, Saha A, Bosma J, Huisman H. Uncertainty-Aware Semi-Supervised Learning for Prostate MRI Zonal Segmentation. arXiv preprint arXiv:2305.05984. 2023 May 10.

22. Yu AC, Mohajer B, Eng J. External validation of deep learning algorithms for radiologic diagnosis: a systematic review. Radiology: Artificial Intelligence. 2022 May 4;4(3):e210064.

23. Widner K, Virmani S, Krause J, Nayar J, Tiwari R, Pedersen ER, Jeji D, Hammel N, Matias Y, Corrado GS, Liu Y. Lessons learned from translating AI from development to deployment in healthcare. Nature Medicine. 2023 May 29:1–3.

24. Clark P, Kim J, Aphinyanaphongs Y. Marketing and US Food and Drug Administration Clearance of Artificial Intelligence and Machine Learning Enabled Software in and as Medical Devices: A Systematic Review. JAMA Network Open. 2023 Jul 3;6(7):e2321792-.

25. Kelly BS, Judge C, Bollard SM, Clifford SM, Healy GM, Aziz A, Mathur P, Islam S, Yeom KW, Lawlor A, Killeen RP. Radiology artificial intelligence: a systematic review and evaluation of methods (RAISE). European radiology. 2022 Nov;32(11):7998–8007. doi.org/10.1007/s00330-022-08784-6

